# A Real-Time Statistical Model for Tracking and Forecasting COVID-19 Deaths, Prevalence and Incidence

**DOI:** 10.1101/2020.05.16.20104430

**Authors:** Jack A. Syage

## Abstract

**Background:** Pandemics do not occur frequently and when they do there is a paucity of forecasting tools that could help drive government responses to mitigate worst outcomes. Here we provide a forecasting model that is based on measurable variables and that strives for simplicity over complexity to obtain stable convergent forecasts of death, prevalence, incidence, and safe days for social easing.

**Methods:** We assume, based on prior pandemic data, that death rate rise and fall approximately follows a Gaussian distribution, which can be asymmetric, which we describe. By taking daily death data for foreign countries and U.S. states and fitting it to an appropriate Gaussian function provides an estimate of where in the cycle a particular population lies. From that time point one can integrate remaining time to obtain a final total death. By also using measured values for the time from infection to recovery or death and a mortality factor, the prevalence (active cases) and incidence (new cases) totals and rate curves can be constructed. It is also possible by setting a downward threshold on prevalence that an estimate of a minimum date to begin relaxing social restrictions may be considered.

**Results:** To demonstrate the model we chose the most severe hot-bed countries and U.S. states as a testbed to evolve and improve our model and to compare with other models. The model can readily be applied to other countries by inputting data from public data bases. We also compare our forecasts to the University of Washington (UW) IHME model and are reassuringly similar yet show less variability on a weekly basis. The sum of squares for error (SSE) for international and U.S. states, respectively, that we track are: 26% and 26% for our model vs. 41% and 46% for the IHME model.

**Conclusions:** Our model appears closest to the UW IHME model; however, there are important differences and while both models forecast many of the same results of interest, each one offers unique benefits that the other does not. We believe that the model reported here excels for its simplicity, which makes the model easy to use.

## INTRODUCTION

Most of the world was caught unprepared by the severity of COVID-19, though retrospectively, there were collective sighs of “we should have seen it coming,” and many did, but these warnings were more opinion than grounded in hard data that the world’s leadership could act on. The number of recorded deaths due to COVID-19 around the world (excluding China) for each month from January to April were 0, 84, 33,878, 189,086, respectively. There is clearly a great need for predictive tools to provide real-time forecasts in order to implement mitigation efforts quicker.

In this paper we describe a real-time statistical model for forecasting COVID-19 deaths, as well as estimating and forecasting the actual number of cases (prevalence) and rate of new cases (incidence). These values are generally greatly understated since they are confirmed by testing that is never in sufficient supply. So, reports of new cases are really reports of new tests of old cases. This model also can set criteria for which some degree of easing of social restrictions can occur on a gradual basis along with check-gate measurements for when these restrictions can be progressively eased. The model can also forecast the consequences of socially easing prematurely. The motivation for this work is to provide predictive tools that are reasonable and devoid of excessive complications that hinder some other models leading to potential inaccuracies and instabilities in the outcome results.

The traditional approach to modeling is epidemiological in nature and seeks to understand known behavior, which for an epidemic, would be after the fact. Conventional epidemiological models include SIR, SEIR and SEIRS (susceptible, exposed, infectious, recovered, susceptible) models^1^,^2^ and are characterized as being mathematically complex and having many variables that are difficult to define.^3, 4^ They are also not well suited to real-time forecasting as new epidemics have their own distinct properties and further it is difficult for them to consider implementation of social restrictions or changes in social behavior due to government easing and/or public rebellion due to economic and social pressures. Our focus here is on a very different approach that uses real-time data to dynamically and statistically forecast important variables of an epidemic as they develop thereby automatically incorporating the epidemic properties and social response as they are embedded in the real-time data.^5^ We do note the development of hybrid epidemiological models for COVID-19 that try to incorporate real-time data, but they still appear to be dependent on many difficult-to-define variables and are very complex.^6,7,8,9,10^ We also note epidemiological models that have used China COVID-19 data to extrapolate forecasts for the rest of the world.^11^

Several real-time statistical models have emerged that specifically address the unique properties and behavior of COVID-19, knowledge that is only learned by following daily trends.^12,13,14,15^ Most of these models are being reported, reviewed and critiqued in the public (digital) forum, justifiably prior to being peer reviewed and published. The rationale for this approach is to provide useful information to help inform policy makers and public implementers on immediate responses that need to be taken, e.g., in deciding when it is prudent to ease social restrictions as they weigh against other considerations, such as the health of the economy. That real-time statistical models often update their forecasts is a strength, not a weakness as long as it is within reason; the adjustments reflect the latest reality, which should not be ignored.

The most widely cited and utilized model for COVID-19 is from the Institute for Health Metrics and Evaluation (IHME) at the University of Washington (UW)^11^ and therefore we consider it a worthy benchmark for our model though we respect other models that attempt to provide similar information. As best as we can determine the IHME model uses the integrated total death curve (S-curve) to determine where on the time progression a population lies in order to integrate to total deaths. Alternatively, we use the death rate curve (1^st^ derivative of the total death curve) as it is more sensitive to detection of the inflection point for peak death rate.^5^ This approach has also been taken by Luo,^16^ however, that model uses new cases (incidence) rather than death, which we consider to be a misleading variable as it is a convolution of actual cases and testing, thereby giving understated values further exacerbated by the testing proportion not being constant (increasing) with time.

## METHOD

On March 14, 2020 we launched a website blog to report our developing model for COVID-19.^5^ We assume, based on prior pandemic data (e.g., COVID-19 in China and 1918 Spanish Flu^17^), that death rate rises and falls approximately following a Gaussian distribution. (We challenge this assumption shortly.) By taking daily death data for foreign countries and U.S. states and fitting it to a Gaussian provides an estimate of where in the cycle a particular population lies. From that time point one can integrate remaining time to obtain a final total death. By also using measured values for the time from infection to recovery or death and a mortality factor, the prevalence (active cases) and incidence (new cases) totals and rate curves can be constructed. It is also possible by setting a downward threshold on prevalence that an estimate of a minimum date to relax social restrictions may be considered.

The general algorithm is to use death rate data and an assumed symmetric or asymmetric Gaussian rise and fall curve to forecast total deaths by estimating where on the Gaussian curve the actual death rate curve lies. Other functions besides a Gaussian could work (and we discuss some of them in the Discussion), but don’t have the mathematical simplicity and the COVID-19 data currently doesn’t provide a compelling reason to consider more complicated functions. We show how this model can accommodate an asymmetric Gaussian or other function, e.g., kinetic curves associated with compartmental approaches. However, a Gaussian is convenient to demonstrate the model and we proceed as such.

The operating equations are as follows:

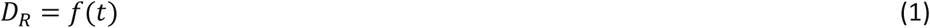

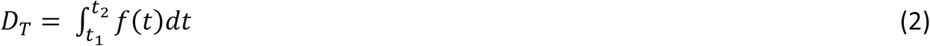

Where *D_R_* and *D_T_* are death rates (e.g., daily) and total deaths, respectively. We first define *f(t)* as a Gaussian function as follows:

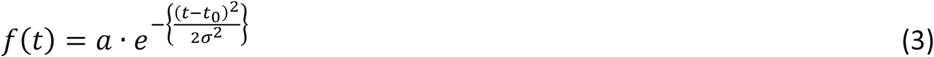

where

> *a* = peak death rate (e.g., daily)
>
> *t* = time relative to the peak death rate time *t*_0_
>
> *σ=* width of the Gaussian where 2.3548σ is the full width at half maximum (FWHM)

Figure 1 shows the basic premise of our algorithm with regard to the relationship of *D_R_* and *D_T_* and how final total deaths are calculated. Figure 1a shows a Gaussian *D_R_* function fitted to data for Italy up to April 2, 2020, which in retrospect is very close to its peak death rate. The integral of Eq. (1) is the non-analytic error function, but which is easily computed (Figure 1b) as follows:

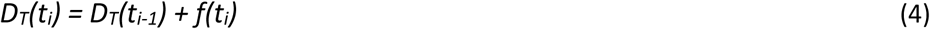

**Figure 1.**
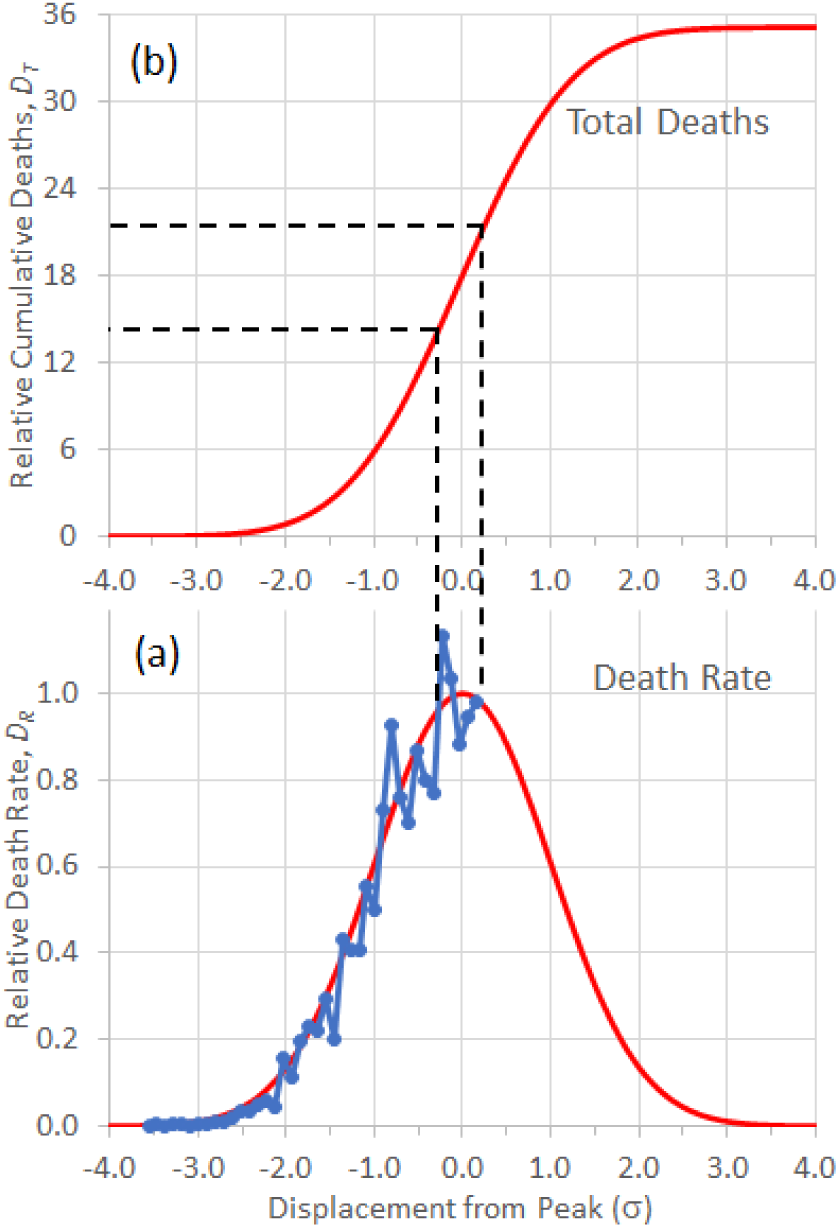
Basic Gaussian model for death rate *D_R_* and cumulative deaths *D_T_*. Data from Italy for 4/2/20 is superimposed by best fit. Abscissa is plotted relative to σ, which for the example shown is assumed to be 2 weeks. The dotted lines show range of uncertainty and how they transpose to cumulative deaths.

Where *i* is the increment of time used for the calculation (e.g., days, weeks). To compute the rate of new cases (incidence), *C_R_*, and the number of current active cases (prevalence), *C_T_*, we make assumptions based on measurable and statistical information. The operating equations are:

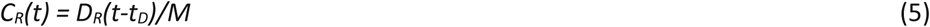

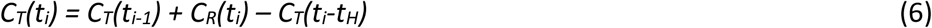

Where *t_D_* and *t_H_* are the times from infection (new case) to death and healed, respectively, and *M* is the mortality rate. The total active cases for *t_i_* is computed similarly for deaths in Eq. (4) except that we subtract the total cases from *t_H_* time ago as being those patient cases who have run their course and either recovered/healed or died.

Figure 2 shows the relationships between *D_R_*, *D_T_*, *C_R_* and *C_T_*. For the purposes of this example we assume that *t_D_* = 2.5 weeks and *t_H_* = 3.5 weeks based on published data for China.^18^ We also assume that the Gaussian rise and fall has a σ= 2 weeks (FWHM = 2.3548 σ), which is consistent with the results for COVID-19 in China and for the 1918 Spanish Flu^17^. Then the current number of deaths on a given day would reflect the number of new cases 2.5 weeks earlier divided by the mortality factor *M* [Eq. (5)].

**Figure 2.**
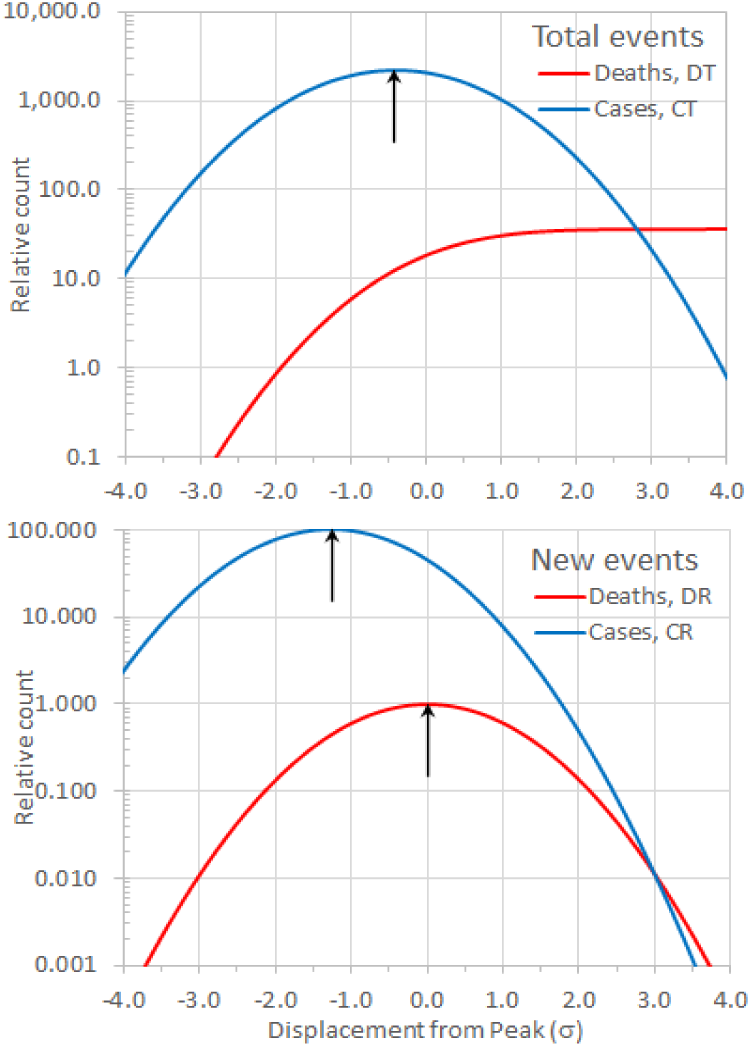
Plots of (a) new deaths and cases, *D_R_* and *C_R_* and (b) total deaths and active cases, *D_T_* and *C_T_*. Time is relative to σ, but in this example we set that to 2 weeks since we are putting real time in for *t_D_* = 2.5 wk and *t_H_* = 3.5 wk. We assume *M* = 1.0%. The arrows denote peaks in the curves. The ordinate is plotted on a log scale in order to cover the large dynamic range of the calculation.

We now summarize some interesting and sometimes non-intuitive observations:

- As assumed by the *t_D_* = 2.5 weeks, the *C_R_* curve peaks 2.5 weeks earlier than the *D_R_* curve.
- The *D_T_* curve asymptotically reaches a final value.
- The *C_T_* curve decreases due to recoveries and peaks about halfway between the *D_R_* and *C_R_* curves.

The above explanation used the simple symmetric Gaussian function and treated many of the variables, e.g., *σ*, *t_D_, t_H_* as constants, based on reported data, in order to provide a basic understanding of the principles to this statistical, real-time model. In the Results section next, we introduce several refinements.

## RESULTS

As an example of the unknowable changing dynamics of COVID-19 and the effects of social response to the disease we show in Figure 3 the death rate plot for Italy that has progressed well past its peak (shown also in Fig. 1). Clearly the ascent and descent are very asymmetric as evidenced by the poor fit to a symmetric Gaussian function on the downslope. Also shown is an asymmetric Gaussian model that incorporates two key refinements:

- An asymmetric Gaussian function based on a non-constant *σ* value.
- The potential to functionalize *σ* to include social factors, e.g., the degree of social distancing and in turn other contributing factors, e.g., population density, mobility, and contract frequency.

**Figure 3.**
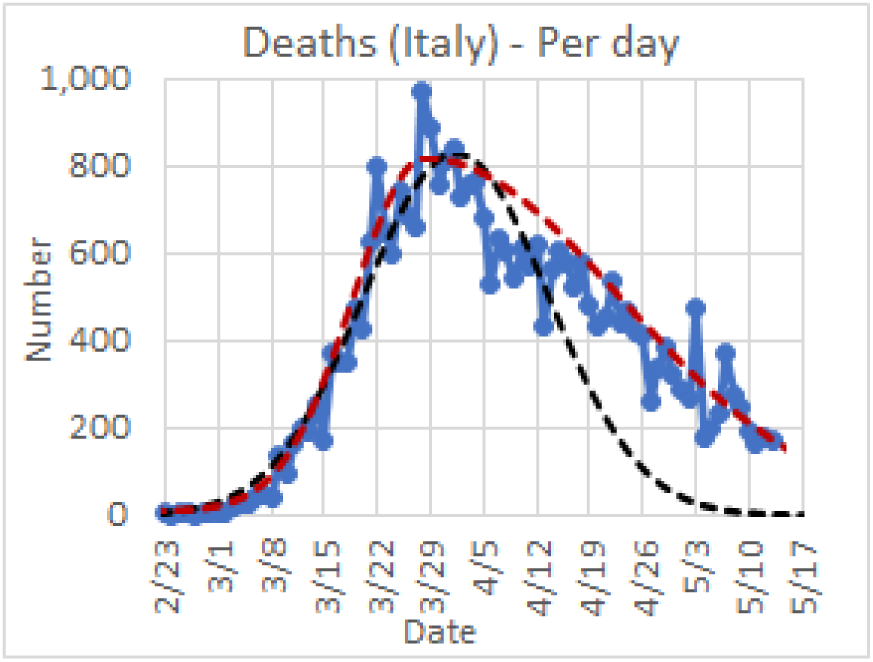
Death rate, *D_R_*, plot for Italy up to 5/12/20 along with a symmetric (black dash) and asymmetric (red dash) Gaussian functional fit. The latter best fit curve corresponds to *σ_a_* and *σ_d_* values of 10 and 24 days, respectively.

### Asymmetric Gaussian Functions

The shape of a Gaussian function is fixed, but its width and asymmetry are embodied by the value *σ*, which is normally a constant value. We consider non-constant treatments for *σ* to try to represent the actual death rate behavior for different global populations.

Figure 4 shows two examples of asymmetric Gaussian functions. Figure 4a shows the case for a two-side Gaussian for which there are separate constant σ values for the ascending (*σ_a_*) and descending (*σ_d_*) sides of the death rate curve, Dr. The example shown is for values of *σ_d_* = 2*σ_a_*, (14 and 28 days if not on a relative abscissa scale), the *σ_d_* value giving a slower descent as representative of the real-time data that we are observing for COVID-19 around the world and in U.S. states. Figure 4b shows the case for a linearly changing *σ*value with a functional form

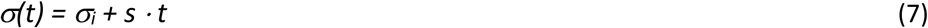

**Figure 4.**
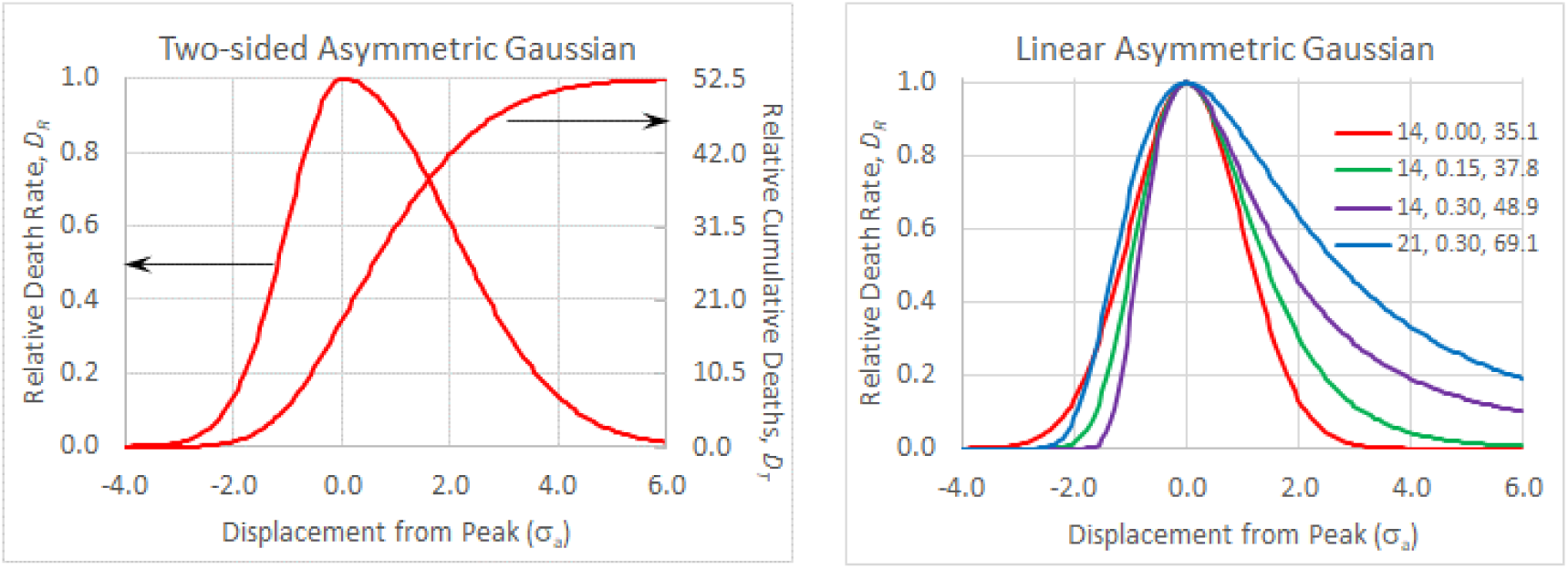
(Left) Ascending and descending width parameters are *σ_a_*/2 *= σ_a_* = 14 days, respectively. (Right) Linearly increasing *σ*value according to Eq. 7 where *σ_i_* and *s* are shown along with the extrapolated final death count, *D_T_*. The abscissa is time in units of *σ_a_* = 14 days.

An important observation is how the shape of the Gaussian influences the final total death count *D_T_*, which is the integral of the *D_R_* death rate curve. For the symmetric Gaussian in Figure 1, the *D_T_* is equal to 35.1 times the peak death rate (in daily units). For the two-sided asymmetric Gaussian in Figure 4a, where the ascending and descending behaviors are described by *σ*and 2*σ*, respectively, the *D_T_* is equal to 52.6 times the peak death rate. In Figure 4b various functions are plotted for various values of *σ_i_* and *s* and the final death count *D_T_* is given. As the descending part of the curve becomes more extended the integral value *D_T_* becomes greater. This has strong implications for populations to continue to practice social isolation to expedite the descent as long as is reasonable weighed against social and economic pressures.

We note that Eq. (7) is just one functional form for *σ(t)* and others may be proposed, which could be utilized to model behavioral changes in the population during the epidemic due to social distancing and other factors. We propose some interesting avenues of development in the Discussion section, but don’t develop this term any further here. It should be noticed though that this linearized (and continuously increasing) function *σ(t)* has the effect of flattening the entrance wing and extending the exit tail of the total curve. While this is less representative of the death rate growth shape, it has minor impact on the integrated death count; however, the elongated and non-converging tail can have large integrated value, which we do not yet have evidence is a good representation. We discuss in the Discussion other functional forms that can represent reported statistic data as well as anticipated social behavior.

### Application to COVID-19

As this manuscript is being prepared in a middle of a pandemic, we can only provide progress on our capability to track and forecast deaths, prevalence, incidence, and safe easing dates. We have been reporting weekly updates for the most serious hotbed countries and U.S. states in a running blog where some details of this model are also presented.^5^ The international data is from the European Center for Disease Prevention and Control (ECDC)^19^ and the U.S. data is obtained from Worldometer.com.^20^ Death rate plots are presented in the Supplemental Material section (Figure S-1) as of May 12, 2020. An example for Italy is given in Figure 3. The results of our calculations and forecasts for international and U.S. populations (as of May 12, 2020) are presented in Table 1.

**Table 1.** Death, prevalence, incidence and easing dates forecasted from the Asymmetric Gaussian model

| Country | Current fatalities | Assumed mortality | Weeks from peak | Calc. prevalence | Calc. Incidence | Final fatalities | Deaths/ million | Prevalence/ million | Easing Date |
| --- | --- | --- | --- | --- | --- | --- | --- | --- | --- |
|  | 5/12/20 |  |  |  |  |  |  |  |  |
| U.S. | 80,684 | 1.0% | 3.3 | 3,547,675 | 83,105 | 114,571 | 346 | 10,718 | 7/4/20 |
| Italy | 30,739 | 1.5% | 6.3 | 190,172 | 2,254 | 32,583 | 539 | 3,145 | 6/9/20 |
| Spain | 26,744 | 1.5% | 5.3 | 171,518 | 1,248 | 28,081 | 601 | 3,668 | 6/4/20 |
| France | 26,643 | 1.5% | 4.3 | 161,634 | 710 | 27,709 | 424 | 2,476 | 5/28/20 |
| U.K. | 32,065 | 1.5% | 3.4 | 546,388 | 7,054 | 37,195 | 548 | 8,049 | 6/15/20 |
| Sweden | 3,256 | 1.0% | 2.9 | 113,309 | 1,693 | 4,037 | 400 | 11,220 | 6/18/20 |
| Iran | 6,685 | 2.0% | 6.1 | 55,318 | 1,003 | 7,554 | 90 | 659 | 5/30/20 |
| S. Korea | 258 | 1.0% | 7.0 | 129 | 1 | 259 | 5 | 3 | Now |
| China | - | 1.0% | 12.0 | - | - | - | - | - | Now |
| WW | 285,936 | 1.5% | 3.9 | 5,398,472 | 106,749 | 351,701 | 45 | 695 |  |
| U.S. States | Current fatalities | Assumed mortality | Weeks from peak | Calc. prevalence | Calc. Incidence | Final fatalities | Deaths/ million | Prevalence/ million | Easing Date |
|  | 5/12/20 |  |  |  |  |  |  |  |  |
| New York | 27,175 | 1.0% | 3.7 | 578,012 | 5,979 | 30,436 | 1,565 | 29,712 | 6/18/20 |
| New Jersey | 9,541 | 1.0% | 2.3 | 446,328 | 8,969 | 13,167 | 1,482 | 50,251 | 7/5/20 |
| Michigan | 4,674 | 1.0% | 3.1 | 110,447 | 1,169 | 5,282 | 529 | 11,059 | 6/13/20 |
| Louisiana | 2,347 | 1.0% | 3.6 | 31,309 | 188 | 2,488 | 535 | 6,735 | 6/4/20 |
| California | 2,876 | 1.0% | 2.9 | 134,827 | 3,566 | 4,343 | 110 | 3,412 | 6/21/20 |
| Washington | 964 | 1.0% | 4.7 | 21,324 | 415 | 1,138 | 149 | 2,800 | 6/17/20 |

We have not yet evaluated an ideal functional form for *σ(t)*, for which Eq. (7) is just one and probably not the best representation, so here we instead use the 2-sided Gaussian model (Figure 4a) and optimize the σ_a_ and σ_d_ values as this function fits well to essentially all death rate data we are following (the σ_a_ and σ_d_ values for each country and U.S. state are presented in Supplemental Material, Table S-1. Notable results in Table I include:

- The U.S. is forecasted to exceed 100,000 total deaths, but the per capita forecast of 346 deaths per million people is below several European countries, though all within a factor of 2x.
- U.S. states tell a different story than the aggregate for the U.S. The forecasted per capita total deaths for New York and New Jersey exceed by many multiples those for the above stated foreign countries. However, we haven’t broken these countries down to states or cities, but on a population basis (N.Y and N.J. at 28.3M vs. 10.1M to 67.9M for all other countries shown except China) one can conclude that N.Y. and N.J. were/are the most deadly large populations on earth.

### Safe Easing of Social Restrictions

Social, economic, and political pressure will ensure that relaxing of social restrictions will occur and probably prematurely. We have developed a protocol to forecast dates for which gradual and measured easing of social restrictions may begin. The easing date is based on the date for which the prevalence per million people drops to 200 or 2 active case per 10,000 people (0.02%). This is a somewhat arbitrary threshold, but seems reasonable based on that approximate value on the front-end was enough to start exponential outbreaks. So, assuming prevalence continues to drop and we don’t revert back to full normal social interactions we should be able to ease while maintaining and even reducing the prevalence. By maintaining the prevalence and assuming a mortality rate of 1% corresponds to one death per million over the course of an infection (about 18 days) and for the U.S. with a population of 331M, would compute to about 18 deaths/day. If we can define a tolerable death rate (perhaps greater than this), then we can back out a different prevalence threshold for social easing. The flatten-the-curve approach is to maintain a death rate sufficiently low to not overload the healthcare system and wait until a vaccine or herd immunity reduces the death rate. However, society may not have the heart to deliberately sustain this level of deaths. This is all weighed against the need to feed economic activity. So ultimately the acceptable death rate comes down to the decision of how many jobs is a death worth. That is a topic outside the scope of this manuscript.

### Importance of Prompt Government Intervention

This model can also estimate the dependence of death rate and total deaths on the date when social distancing is implemented. We all recall how frightening it was to be seeing exponential growth in deaths and reading about its doubling every so many days. These deaths of course reflected exponential growth of new cases about 2.5 weeks earlier representing the date of the infection that led to these deaths. This alone drives home the point that swift action to reduce contact exposure could have huge significance on reducing overall deaths, but we can also semi-quantify this reality.

Figure 5 presents a pedagogical illustration, but one modeled somewhat on the U.S. response. Here we forecast the difference in the occurrence of death based on starting social distancing at time points two weeks apart (3/4/20 vs. 3/18/20). For the assumed values (see figure caption), a 2-week delay in implementing social distancing has a consequence of a 5x increase in deaths. This model is very simple since it assumes a symmetric Gaussian, but the qualitative conclusions are nonetheless revealing and sobering.

**Figure 5.**
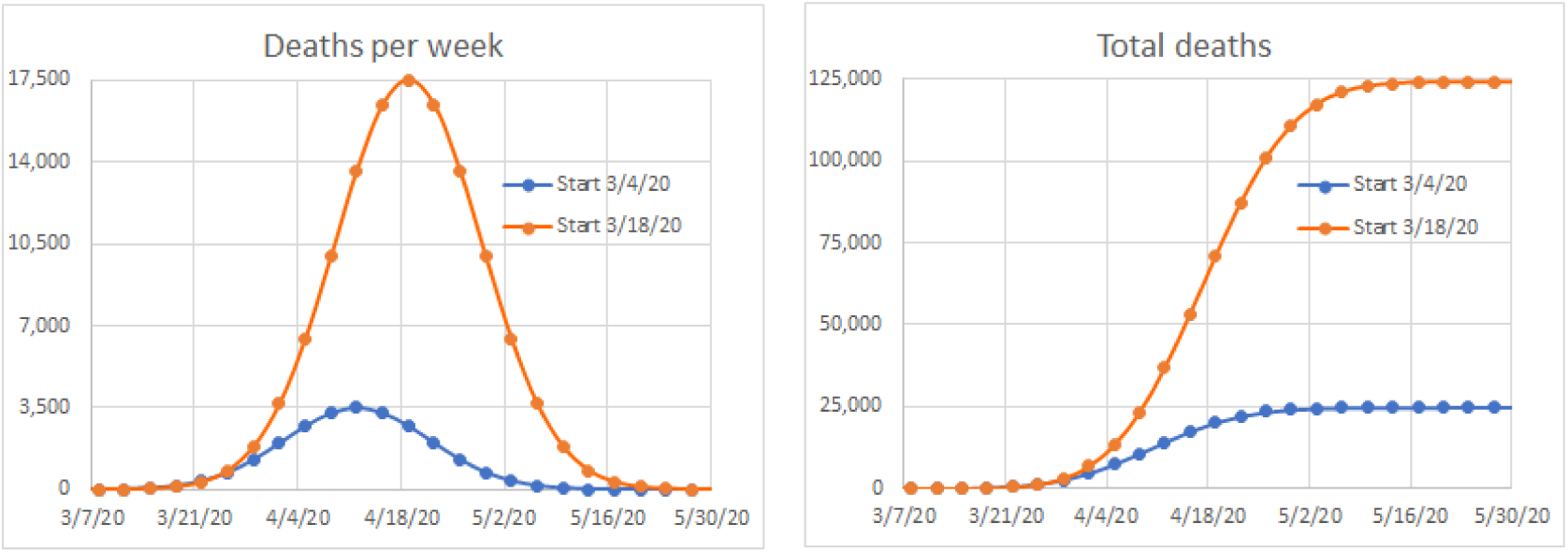
Gaussian model forecast for death rate and total deaths for social distancing implementation on 3/4/20 vs. 3/18/20 for assumptions representative of the U.S. population. σ = 14 weeks.

### Comparison to Other Models

We do not compare ourselves to epidemiology models as explained in the Introduction because the purpose of our development is very different, namely providing forecasting information based on latest data, so there will necessarily be changes in the forecasts as new data is reported. The former approach is too rigid to be a forecasting tool, but essential for understanding the properties of an epidemic once all the facts are known. Real-time statistical forecasting models are an invention of their times as each pandemic, rare in themselves, are different and therefore a model has to react to these differences. For COVID-19 the most widely cited and utilized model is from the Institute for Health Metrics and Evaluation (IHME) at the University of Washington (UW)^12^ and therefore we consider it a worthy benchmark for our model. We have been reporting our forecasts relative to the IHME forecasts on a weekly basis.^5^

Figure 6 shows the total death forecasts for hotbed international countries by the two models and shows that there is more similarity than differences. (Results for U.S. states are given in Figure S-2 in Supplemental Material.) Still there are differences and that underlines the need to endorse multiple models, even if similar, to provide a range of outcomes representing different, even if subtle, algorithms and assumptions. A true comparison awaits the end of this wave of the pandemic or picking a target date as IHME has for August 4, 2020. Our forecasts extrapolate to the end of the Gaussian tail, but there is very little integration value beyond August 4, 2020 so our comparison is suitable. At this present time, one figure of merit for the stability of a model is how much variation or volatility it has from one time point to the next. In Figure 7 we plot the sum of squares for error (SSE) measure of variability for the weekly forecasts presented in Figure 6. We caution that variability is necessary because it responds to real-time changes and results, but if excessive can also be detrimental for policy and planning purposes.

**Figure 6.**
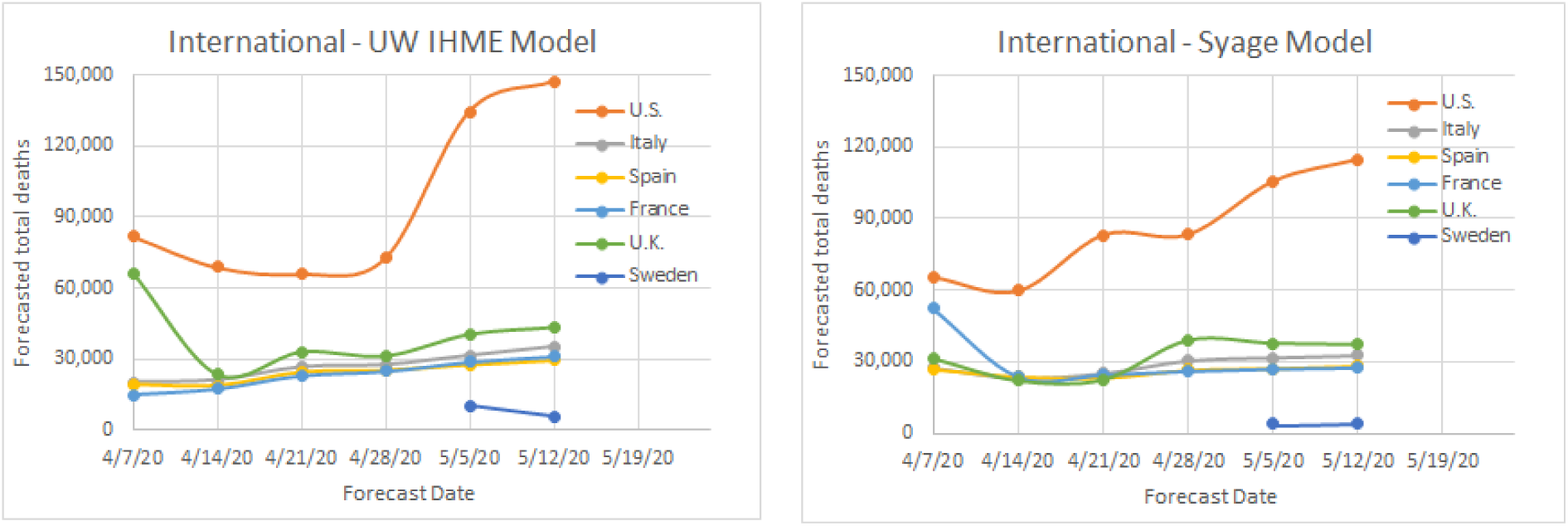
Plots of total death forecasts by the IHME and Syage models for hotbed countries.

**Figure 7.**
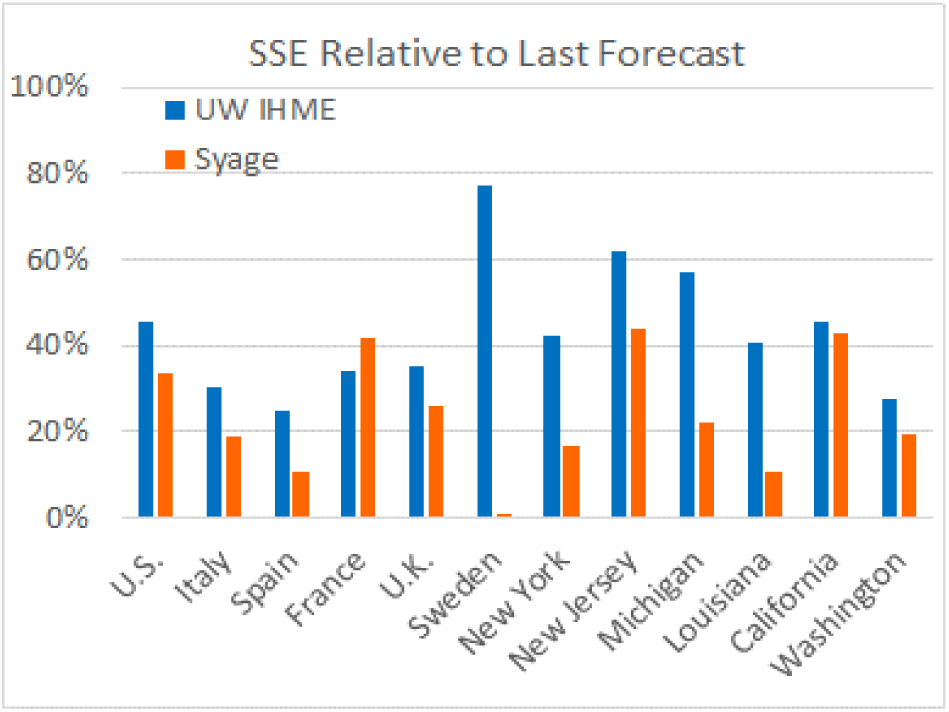
Sum of squares for error (SSE) calculated as:

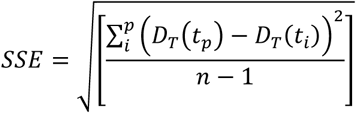 Where *t_p_* is present time and *t_i_* are previous time periods. Updated to 5/12/20.

By this SSE measure the IHME model forecasts have varied greater from week to week than the present model for all but one of the cases shown (France). If averaged for the international and U.S. states, respectively, that we track the SSE’s are: 26% and 26% for our model vs. 41% and 46% for the IHME model. At present we do not see a penalty to the present model’s relative stability, but time will tell.

## DISCUSSION

The above results describe a forecasting model that is simple in principle, but with capability to address more subtle dynamics of the on-going COVID-19 pandemic. We now discuss some of these present and future refinements.

Clearly the Gaussian model is simple and instructive for informing how to structure a data-driven statistical predictive model. However, it is now clear that whereas it is very descriptive of the growth phase of the epidemic, it does not represent the recovery side very well, at least not in its symmetric construction. We have shown that simple asymmetric Gaussian functions so far appear to fit well to the recovery side. We are satisfied with the asymmetric function with an ascent and descent sigma values, *σ_a_* and *σ_d_* (Figure 4 left), even though it somewhat violates our sense of an abruptly changing sigma value. Alternatively, the linear sigma functional construct (Eq. (7) and Fig. 4 right) had detriments in the wings and the tails.

We have explored other functions. Gamma and Poisson distributions have convenient and computable asymmetric shapes that could be fitted to real-time data, but we do not see an advantage over using an asymmetric Gaussian with a functionalized sigma *σ(t)*. Many manifestations of SIR models have the basics of bi-exponential functionality. However, the bi-exponential growth phase does not show an exponential rise or an inflection point as it progresses to a peak rate, but rather has a progressively decreasing rate from inception so we dismiss this functional form. Of course, any of these functions can be modified to build in the dynamical characteristics of a true epidemic response, but the math becomes very complex.

We are intrigued by the use of asymmetric Gaussian functions, particularly by functionalizing the sigma value *σ(t)* beyond the two-valued and the linear constructs discussed above. In future work we plan to explore how to functionalize *σ(t)* in such a way that it can incorporate social isolation time dependences. This may include a sum of displaced Gaussians to represent different waves and outbreaks.

It is also useful to consider other means to identify the approach to death rate and other peaks. We became aware of an interesting log-log plot that we have applied to death rate *D_R_* vs. total deaths, *D_T_*, which is shown in Figure 8.^21^

**Figure 8.**
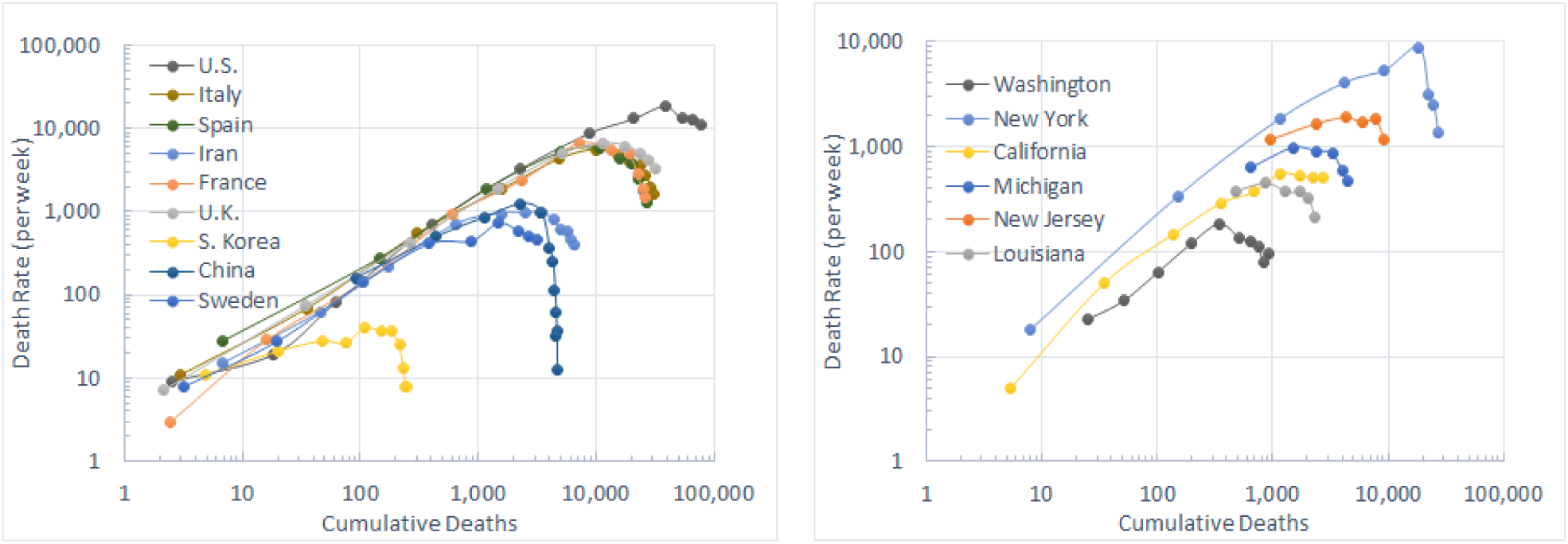
Log-Log plot showing downward deviations representing exit from exponential growth. Each point represents a time interval of one week (updated to week ending 5/13/20).

Figure 8 shows downward deviations representing exit from exponential behavior. This is followed soon by peaking and downward recovery, which are not evident in the log-log plots, but are clearly evident in the rate curves, e.g., Figures 1a and 3. However, the rate curves do not show exiting from exponential growth very well so the combination of these plots provides a better estimation of the progression of an epidemic and where the current death rate lies on the fitting function *f(t)*. Exiting from exponential growth generally occurs about 2 weeks before reaching peak death rate so this information.

Finally, we address the uncertainty of defining a mortality factor, *M*. That can never be known for sure until after the pandemic has run its course though there are ways to infer it, which is necessary for improving accuracy for calculating and forecasting true prevalence, not just confirmed tested cases. *M* will also obviously vary depending on the demographics of the populations (e.g., age distribution, healthcare system) and even change for a particular population due to changing dynamics such as crossing the threshold for overwhelming the healthcare system both on the way and down the epidemic curve. We have made adjustments to our *M* value, particularly for Spain and Italy varying it between 2.0 to 1.5%. Most determinations in the literature are placing this value in the 1.0-1.5% range, ^12,15,18^ which is what we generally use as noted in Table 1.

## CONCLUSION

In this manuscript we describe a predictive model that is based on measurable variables and that strives for simplicity over complexity to obtain stable convergent forecasts of death, prevalence, incidence, and safe days for social easing. This real-time, data-driven statistical approach is to be viewed as an alternative and not competitive model to epidemiological models (e.g., SIR, SEIR, etc.) that attempt to forecast epidemic behavior from collective knowledge and data from prior epidemics. Consequently, real-time statistical models in general will revise their forecasts as new data is reported.

We compare our forecasts to the venerable UW IHME statistical model, which apparently has similar algorithmic constructs as this model as evidenced by the similar outcomes, but differ in key ways that make both models (and others of course) essential for testing different model assumptions, none of which can be truly validated until the properties of an epidemic are known at the end. However, the model described here, up to this point in time (March 12, 2020), displays less forecast variability and may even be a little ahead of trends when compared to the UW IHME model. There does not appear to be a penalty to this greater forecasting stability, though only time will tell. Forecast stability is vital for informing public policy and response.

## Data Availability

All data will be made available upon request and eventually will be placed in a publicly accessible depository.

https://syage-covid19-assessment.com/

## SUPPLEMENTAL MATERIAL

**Figure S-1.**
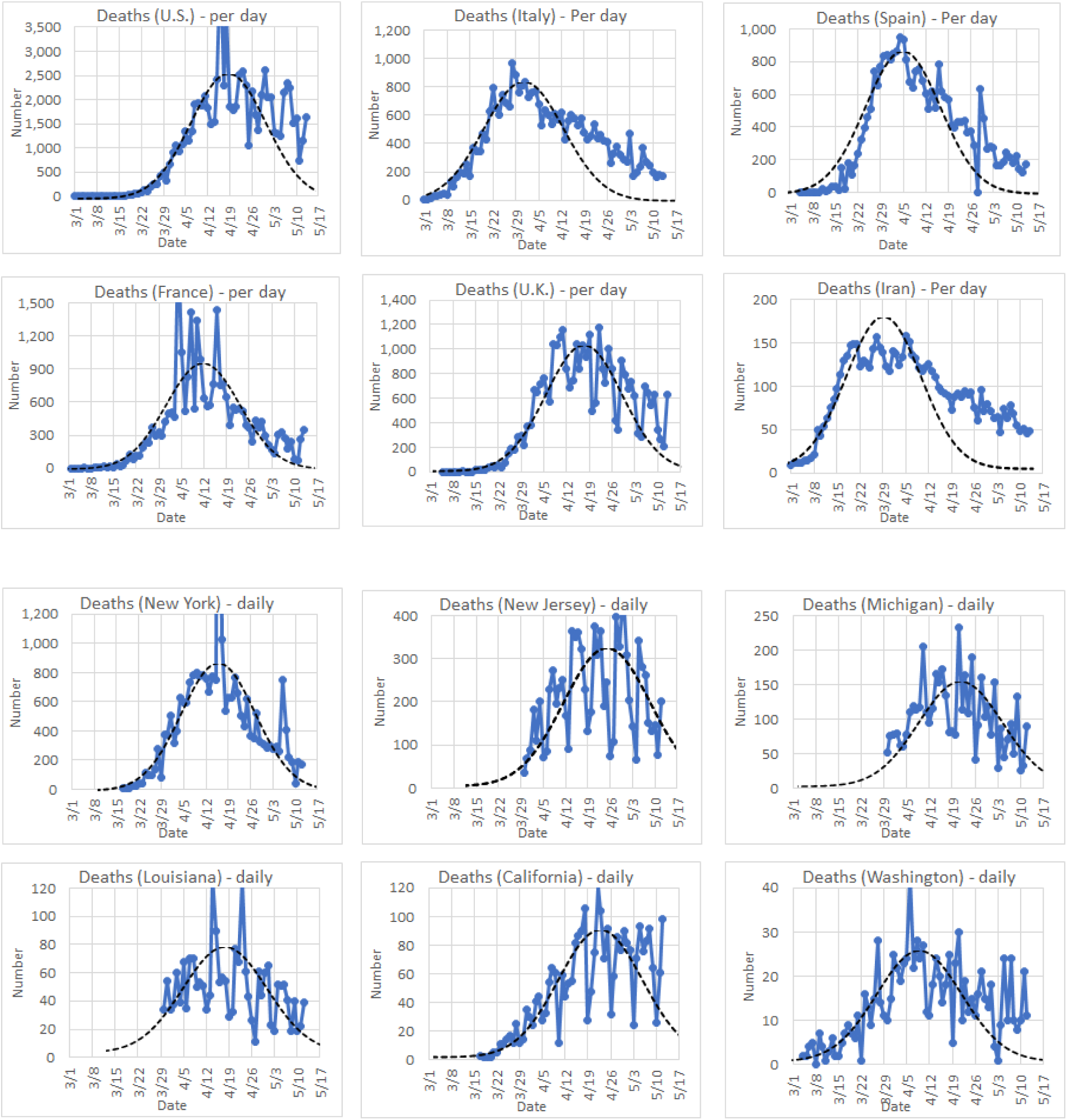
Death rate plots for hotbed countries and U.S. States. The symmetric Gaussian function (dashed curve) is presented for visualization of reasonable fitness on the rising side and less so on the falling side of the death rate plots.

**Table S-1.** Tabulation of the asymmetric Gaussian model fitting parameters for May 12, 2020.

| Country | $\sigma_a$ | $\sigma_b$ | $D_R$ Peak Date | U.S. State | $\sigma_a$ | $\sigma_b$ | $D_R$ Peak Date |
| --- | --- | --- | --- | --- | --- | --- | --- |
| U.S. | 10 | 28 | 4/19/20 | New York | 10 | 19 | 4/16/20 |
| Italy | 10 | 25 | 3/29/20 | New Jersey | 14 | 22 | 4/26/20 |
| Spain | 7 | 20 | 4/5/20 | Michigan | 14 | 18 | 4/20/20 |
| France | 10 | 16 | 4/12/20 | Louisiana | 14 | 16 | 4/17/20 |
| U.K. | 12 | 20 | 4/18/20 | California | 14 | 30 | 4/22/20 |
| Sweden | 12 | 20 | 4/22/20 | Washington | 14 | 28 | 4/9/20 |
| Iran | 8 | 30 | 3/30/20 |  |  |  |  |
| S. Korea | 14 | 16 | 3/24/20 |  |  |  |  |
| China | 10 | 13 | 2/18/20 |  |  |  |  |
| WW | 14 | 26 | 4/15/20 |  |  |  |  |

**Figure S-2.**
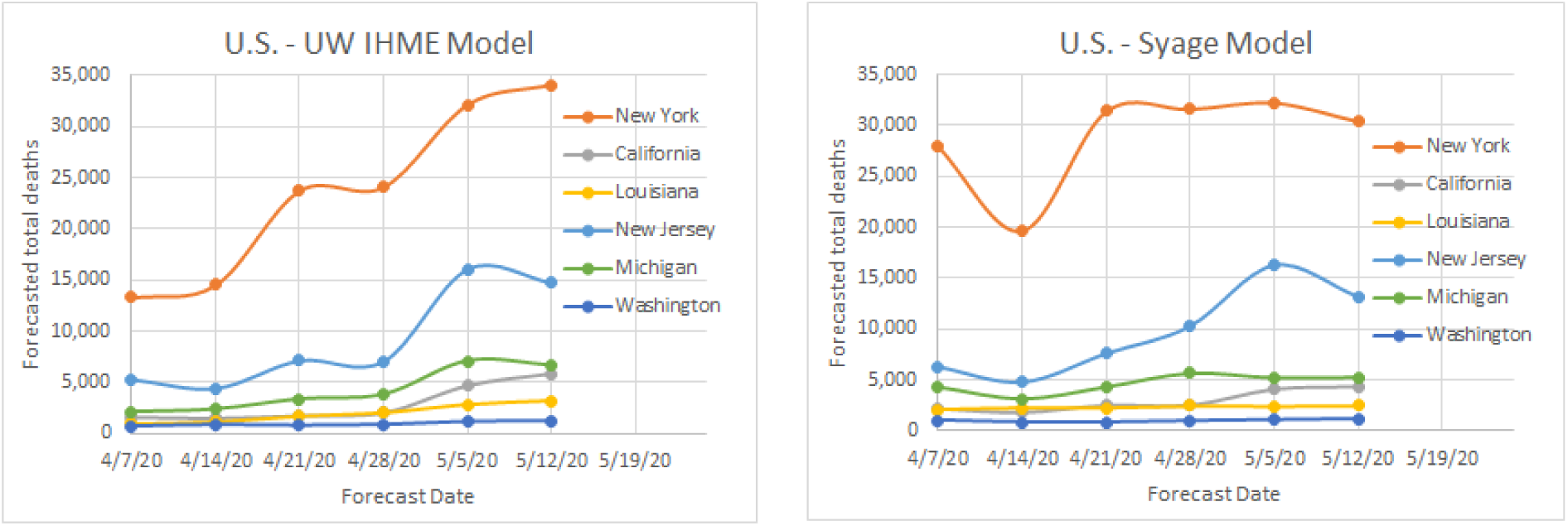
Plots of total death forecasts by the IHME and Syage models for hotbed U.S. states.

